# Use of genetic correlations to examine selection bias

**DOI:** 10.1101/2023.04.04.23288120

**Authors:** Chin Yang Shapland, Apostolos Gkatzionis, Gibran Hemani, Kate Tilling

## Abstract

Observational studies are rarely representative of their target population, because there are known and unknown factors that affect an individual’s choice to participate (known as the selection mechanism). Selection can cause bias in a given analysis, if the outcome is related to selection (conditional on the other variables in the model). However, the selection mechanism usually cannot be detected from the observed data if we have no data on the non-selected sample - for example, when the selected sample is participants in a research study. Here, we develop methods to examine the selection mechanism by comparing correlations among variables in the selected sample to those expected under no selection. We examine the use of four hypothesis tests to identify induced associations between genetic variants in the selected sample. We evaluate these approaches with Monte Carlo simulations. Finally, these approaches are demonstrated with an applied example, using data from UK Biobank (UKBB), with alcohol intake as exposure to test the presence of selection bias. The proposed tests have identified selection due to alcohol intake into UKBB, and the subsample of individuals with weekly alcohol intake. Analyses in UKBB with alcohol consumption as exposure or outcome may be biased by this selection.

## 1 Introduction

In statistical inference, we usually assume the sample under analysis is representative (i.e. a random subsample) of the target population. This assumption could be violated by participants being non-randomly selected into the study sample. This can occur when measured and unmeasured factors affect initial study enrollment or loss to follow up, or when the analysis is limited to a selected group (e.g. only those with no disease at baseline). The distortion of the parameter estimate between the analysis sample and the true value in the target population, is known as “selection bias” (Rothman et al., 2008).

In complete case analysis, i.e. analysis only using the individuals who have complete data, selection causes bias in the estimated exposure regression coefficient if selection is related to both exposure and outcome (or just outcome for linear regression) (J. W. Bartlett et al., 2015; Hughes, Heron, et al., 2019). In Mendelian randomisation (MR), a popular method for estimating causal effects using genetic variants that yields estimates unaffected by unmeasured confounding of exposure and outcome, selection bias may occur if selection is related to exposure or outcome (Hughes, Davies, et al., 2019). Therefore in order to examine the likelihood of selection bias in a given analysis, we need to examine whether exposure or outcome could cause selection. It is possible to examine the association of measured variables with selection, if these variables have also been measured in non-selected individuals. If the selection is of participants into a study, then information on non-participants is usually non-existent or minimal, and thus it is hard to examine which variables are related to selection. However, causes of exposure (or outcome) that are independent in the target population would be dependent if exposure (or outcome) causes selection. Thus correlations between these causes of exposure/outcome observed within the sample would indicate that it was plausible that exposure/outcome caused selection, and that therefore the exposure-outcome analysis may be biased.

Most studies are prone to non-random selection, for example one of the largest cohort studies, UK Biobank (UKBB), has been shown to differ from the UK population in various characteristics (Fry et al., 2017). UKBB is often used for genetic analyses, and in particular MR, and some analyses have been found to be biased by selection (Munafò et al., 2018). For example, the association between alcohol intake and cardiovascular disease has been found to be under-estimated in UKBB (Stamatakis et al., 2021). One aspect of genetic studies that can help with the detection of selection bias is that different unlinked genetic variants (i.e. those on different chromosomes) should (after quality control) be independent (Pirastu et al., 2021). In other words, genetic variants should be independent in the target population. If they are found to be correlated in a study sample, one reason for this would be selection bias. More specifically, shown by Figure 1, suppose we have a population of individuals, *i* = 1, .., *N*. Let the variable *X* be caused by *p* independent genetic variants *G*_*j*_ (*j* = 1, …, *p*). We define *S* = 1 if a participant is selected into the study and *S* = 0 if they are not. Conditioning on *S* induces associations between all causes of *S*, hence the genetic variants *G*_*j*_ will be correlated in the selected sample.

**Figure 1:**
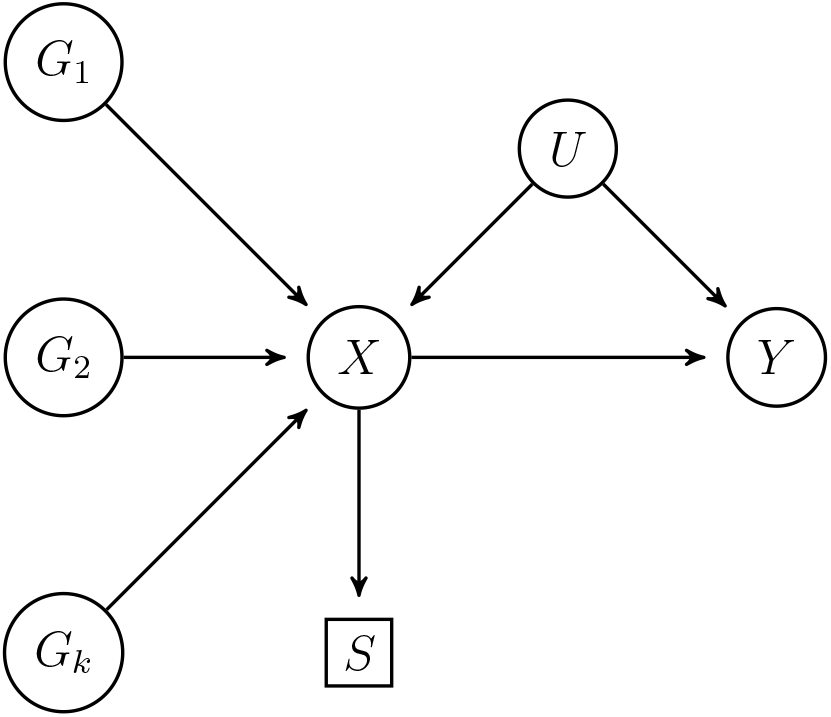
Causal diagrams representing the hypothesized relationship between *p* genetic instruments, *G*_*j*_ (*j* = 1, …, *p*), exposure (*X*), outcome (*Y*), selection variable (*S*), and all unmeasured variables (*U*) which confound *X* and *Y*.

Therefore, to detect selection bias we can examine associations between variables that should be independent in the target population (e.g. genetic variants causing a given phenotype). One approach would be to examine every pairwise correlation coefficient, but this would require many tests to be conducted, and thus correction for multiple testing (Larzelere & Mulaik, 1977). Therefore, we have focused on approaches that test homogeneity in correlation/covariance matrices directly. M. S. Bartlett, 1951, Jennrich, 1970 and Steiger, 1980 proposed test statistics for comparing correlation matrices. (Box, 1949) compares covariance matrices. Our aim is to examine the use of these four hypothesis tests to examine induced associations between genetic variants in a selected sample. We consider two scenarios for identifying selection; ‘one sample’ or ‘two samples’. The former examines the evidence for a single sample not being a random subsample of the target population, by comparing the observed correlation matrix to that expected in the target population (the identity matrix). The latter examines the evidence for two different samples not being from the same population. This is particularly relevant in the case of two-sample MR, which derives causal effect estimates with summary statistics obtained from two separate samples - one provides the SNP-exposure associations and the other provides the SNP-outcome associations. A key assumption is that both samples come from the same underlying population (Bowden et al., 2017). With full individual patient data on both samples one could assess which factors were associated with selection into each sample - but with only summary data, as are usually available, this cannot be done. Comparison of correlations between genetic factors could instead be used to assess the plausibility of this assumption.

We begin with a motivating example using a MR study from UK Biobank (UKBB). Section 2 will introduce the different hypothesis testing approaches. Section 3 will describe the method of simulation using the ADEMP framework (Morris et al., 2019) to evaluate these approaches. Section 4 will give the results of Monte Carlo simulations. Finally these approaches will be applied to the motivating example to test for the presence of selection bias.

### 1.1 Motivating Example: alcohol consumption in UK Biobank

A recent MR study showed that alcohol consumption has a causal effect on risk of having stroke (Larsson et al., 2020). The instruments used were single nucleotide polymorphisms (SNP) hits from a Genome-wide association study (GWAS) on alcohol use (Liu et al., 2019), in which the authors defined “alcohol use” as weekly alcohol intake. However in UKBB, an individual’s weekly alcohol intake is only measured if the participant indicated that their alcohol consumption was “more often than once or twice a week”. This meant that the complete case sample excluded the 35% of total sample of UKBB (total N=501,532) who did not have weekly alcohol intake measured. We hypothesise that in the selected sample (those with alcohol intake measured), the SNPs associated with alcohol frequency will be correlated with each other. This means that in UKBB an analysis using alcohol use as the outcome (including the first stage of an MR analysis with alcohol use as the exposure) would be prone to selection bias.

## 2 Methods

We first describe the assumed model and define the correlation and covariance matrices for the one sample case, then for two sample case.

In the one sample case, suppose that we have *p* variables, **V** = (*V*_1_, …, *V*_*p*_)^*T*^ following some distribution with mean *µ* = (*µ*_1_, …, *µ*_*p*_) and covariance matrix **Σ** = (*σ*_*i,j*_)_*p*×*p*_. For a sample of *n* individuals, the sample mean is 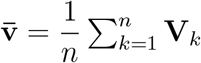 and sample covariance matrix 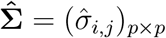 is defined as

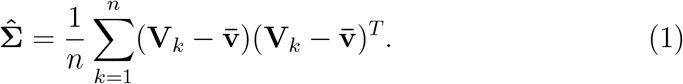

The sample correlation matrix is defined as 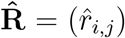, and is estimated from the sample covariance matrix;

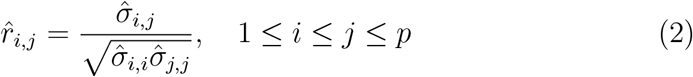

In the two samples case, we have two independent random samples, **V**_1_, …, **V**_*n*1_ and **D**_1_, …, **D**_*n*2_, both are i.i.d. from a *p*-variate distribution with *µ*_**1**_ = (*µ*_1,1_, …, *µ*_*p*,1_)^*T*^ and covariance matrix **Σ**_1_ = (*σ*_*i,j*,1_)_*p*×*p*_, and with *µ*_**2**_ = (*µ*_1,2_, …, *µ*_*p*,2_)^*T*^ and **Σ**_2_ = (*σ*_*i,j*,2_)_*p*×*p*_ respectively. Then we define the sample means by 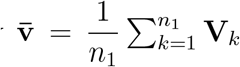 and 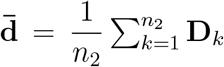 and the sample covariance matrices 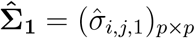 and 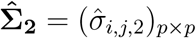 by

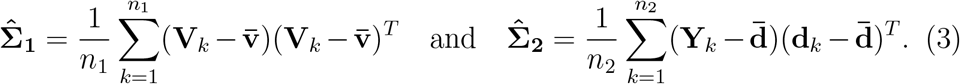

Let **R**_**l**_ = (*r*_*i,j,l*_) be the correlation matrices of *l* sample, where *l* = 1, 2. Then the sample correlation matrices are 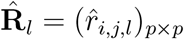 with

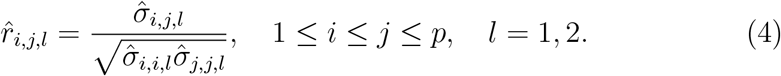

If we have external information that the correlation matrix in the target population would be the identity matrix (e.g. if the **V** = (*V*_1_, …, *V*_*p*_)^*T*^ are genetic variants, which are expected to be independent after quality control) then the null hypothesis for testing for correlation of the **V** = (*V*_1_, …, *V*_*p*_)^*T*^ in the one sample case;

I. *H*_0_ : **R** = **I** For a comparison of the associations between two samples, the null hypothesis is;
II. *H*_0_: **R**_**1**_ = **R**_**2**_ or *H*_0_ : **Σ**_**1**_ = **Σ**_**2**_

where *I* is the identity matrix. *H*_0_ : **R** = **I** and *H*_0_ : **Σ** = *σ*^2^**I** are essentially equivalent as they both test whether the off-diagonal of the matrix is zero. This one sample equivalence of the correlation matrix to the identity matrix is formally known as the “Identity Hypothesis”. For (II), we assume that there are no overlapping individuals between the two samples. As hypothesis testing (I) requires one sample and (II) requires two, we hereafter refer (I) and (II) as hypothesis testing for one-sample and two-sample respectively.

All of the following tests are *χ*^2^ tests (or likelihood ratio test) and commonly used for low-dimensional settings i.e. *p < n* (Cai, 2017).

### 2.1 Testing the identity hypothesis for one sample

#### 2.1.1 Bartlett test

(M. S. Bartlett, 1951) has proposed the following test statistic;

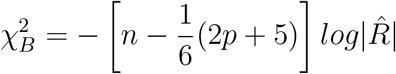

which is assumed to be distributed as a *χ*^2^ with *p*(*p*−1)/2 degrees of freedom.

#### 2.1.2 Jennrich test

The Jennrich, 1970 test defines 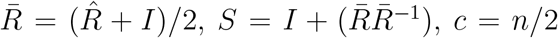 and 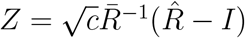. Then

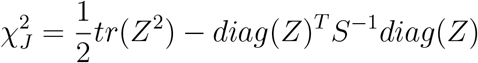

with *p*(*p*− 1)/2 degrees of freedom. The first term on the right is testing the equality of the covariance matrices, and second term is a correction term for testing correlation matrices.

#### 2.1.3 Steiger test

Steiger, 1980 described the test statistic for one sample as,

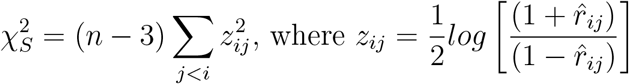

with *p*(*p*− 1)/2 degrees of freedom. For small samples, this test statistic performs better than Jennrich’s, as the Fisher’s *r*-to-*z* transformation ensures the correlation coefficients are normally distributed (Neill & Dunn, 1975).

### 2.2 Testing the equality of correlation/covariance matrices from two samples

#### 2.2.1 Box’s M test

Box, 1949 proposed the M test;

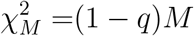

with degrees of freedom *p*(*p* + 1)/2. M is the test statistic and q is the scale factor that ensures the test statistic is asymptotically distributed as *χ*^2^ even with small samples. *M* and *q* are derived from *v*_*l*_ = *n*_*l*_ − 1, 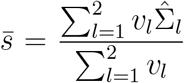, resulting in;

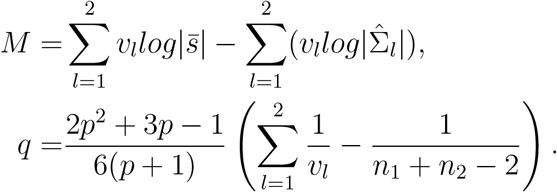

Note that Box, 1949’s M test can test the equivalence of multiple covariance matrices (defined by *l*), however we have simplified the test statistics to test the equivalence of two covariance matrices.

#### 2.2.2 Jennrich test

Jennrich, 1970 described testing the off-diagonal of the difference between two correlation matrices (*R*_1_ −*R*_2_) against the zero matrix. Let 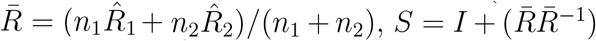, *c* = *n*_1_*n*_2_/(*n*_1_ + *n*_2_) and 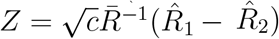. Then

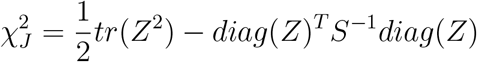

with *p*(*p* − 1)/2 degrees of freedom. As with testing the identity hypothesis for one sample, the first term on the right is testing the equality of the covariance matrices, and second term is a correction term for testing correlation matrices. If the two samples are independently drawn from the same underlying population, then as *n*_1_, *n*_2_ *→ ∞*, 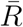 will tend to the population correlation matrix R and the difference between *R*_1_ and *R*_2_ will tend to zero (a matrix with all its elements equal to zero).

#### 2.2.3 Steiger test

For testing the difference between two sample correlation matrices, the Steiger, 1980 test simply becomes

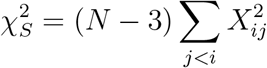

with *p*(*p* − 1)/2 degrees of freedom, where 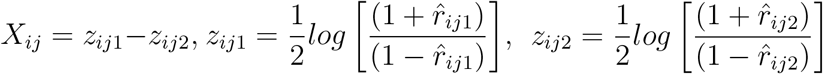, and 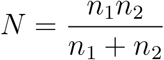.

## 3 Simulation study plan

This section follows the ADEMP framework (Morris et al., 2019).

### 3.1 Aims

The simulation study aims to compare the performance (in terms of type-I and type-II error) of Bartlett, Jennrich, Steiger and Box’s M test for testing the correlation/covariance matrix of a set of genetic variants (SNPs), varying: sample size (*N*); amount of variance in exposure (*X*) explained by these SNPs 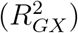, and number of SNPs (*p*) with various minor allele frequencies (MAF). The simulations use ranges for 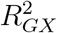 and *N* that are typically seen in Mendelian randomisation studies.

### 3.2 Data-generating mechanisms

The genotypes for SNP *k* with specified MAF, *f*_*k*_, are created first by simulating a latent variable 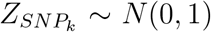, then coding the genotype as 0 if 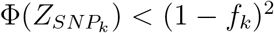, 1 if 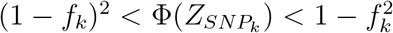, and 2 otherwise, where Φ() is the standard normal integral. The MAFs from all the SNPs are simulated from uniform distribution of *Unif*(0.1, 0.5).

Each simulated dataset has information on *p* SNPs (*G*_*k*_) and phenotype (*X*) on *N* individuals. *X* is normally distributed and 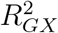 is the proportion of variance of *X* explained by the *p* SNPs. The following describes their relationship;

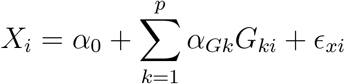

where *ϵ*_*xi*_ is independent random errors of *X* distributed as *N*(0, 1), *i* = 1, …, *N*. As *G*s explain 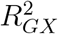 of the variance in *X*, the remaining 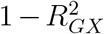 is explained by *ϵ*_*xi*_ and their regression coefficients are calculated accordingly.

Two types of selection (i.e. selection into the study sample, by having X observed) will be simulated:

- selection completely at random (SCAR);
- selection at random, conditional on X (SAR).

Within each simulated dataset, 60% of the *N* individuals are selected (i.e. have exposure data observed). For SCAR, 60% of individuals were randomly selected. For SAR, the following gives the probability of participants being selected (Hughes, Davies, et al., 2019):

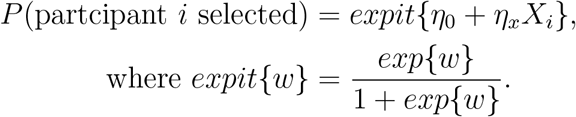

The parameters *η*_0_ and *η*_*x*_ are chosen to give mean probability of selection of 0.6 and standard deviation of 0.2, which reflects 60% of *N* individual having exposure data.

Table 1 gives the summary of varied and default parameters for different scenarios. Each scenario will be simulated 1,000 times.

**Table 1:** Summary of simulation scenarios. *p*, number of *G*s; *N*, sample size; 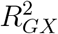, total variance explained by *G*s on X.

| Scenario | $p$ | $R_{GX}^2$ | $N$ |
| --- | --- | --- | --- |
| **One sample testing** |  |  |  |
| 1 | 50 | 0.45 | 2000-10000 |
| 2 | 10-90 | 0.45 | 8000 |
| 3 | 50 | 0.05-0.45 | 8000 |
| 4 | 50 | 0.05 | 10000-400000 |
| 5 | 10-90 | 0.05 | 8000 |
| **Two sample testing** |  |  |  |
| 6 | 50 | 0.45 | 5000-20000 |
| 7 | 10-90 | 0.45 | 10000 |
| 8 | 50 | 0.05-0.45 | 10000 |
| 9 | 50 | 0.05 | 10000-400000 |
| 10 | 10-90 | 0.05 | 10000 |

### 3.3 Estimands

Our estimand is the p-value from testing the null hypotheses described in Section 2.

### 3.4 Methods

In the one-sample case the correlation matrix from selected individuals (those with exposure observed) was compared to the identity matrix using the following methods (as described in Section 2):

1. Bartlett;
2. Jennrich;
3. Steiger.

For comparing correlation/covariances from two samples, the correlation/covariance matrix for the unselected sample (individuals without exposure observed) is compared to the correlation/covariance matrix from the selected sample (individuals with exposure observed) using the following methods (detailed in Section 2):

1. Box’s M;
2. Jennrich;
3. Steiger.

### 3.5 Performance measures

The performance is measured by the proportion of the simulated datasets for which the p-value for the specified test was *<* 0.05. Where there is no selection (SCAR), this should be 0.05, i.e. the nominal type-I error rate. Where there is selection based on exposure (SAR), an ideal test would have a high proportion of tests giving a low p-value.

## 4 Results

### 4.1 Testing the identity hypothesis using one sample

When testing the null hypothesis that a correlation/covariance matrix is equal to the identity matrix, Steiger, Jennrich and Bartlett tests had the correct nominal 5% Type I error rate (T1E) under SCAR for all scenarios (Table 2). Under SAR, for all three methods, the proportion of tests correctly rejecting the null hypothesis increases with sample size and variance explained, and decreases with number of *G*s. All methods correctly reject the null hypothesis in all 1,000 simulated datasets when the sample size and variance explained are large, and number of *G*s are small.

**Table 2:**
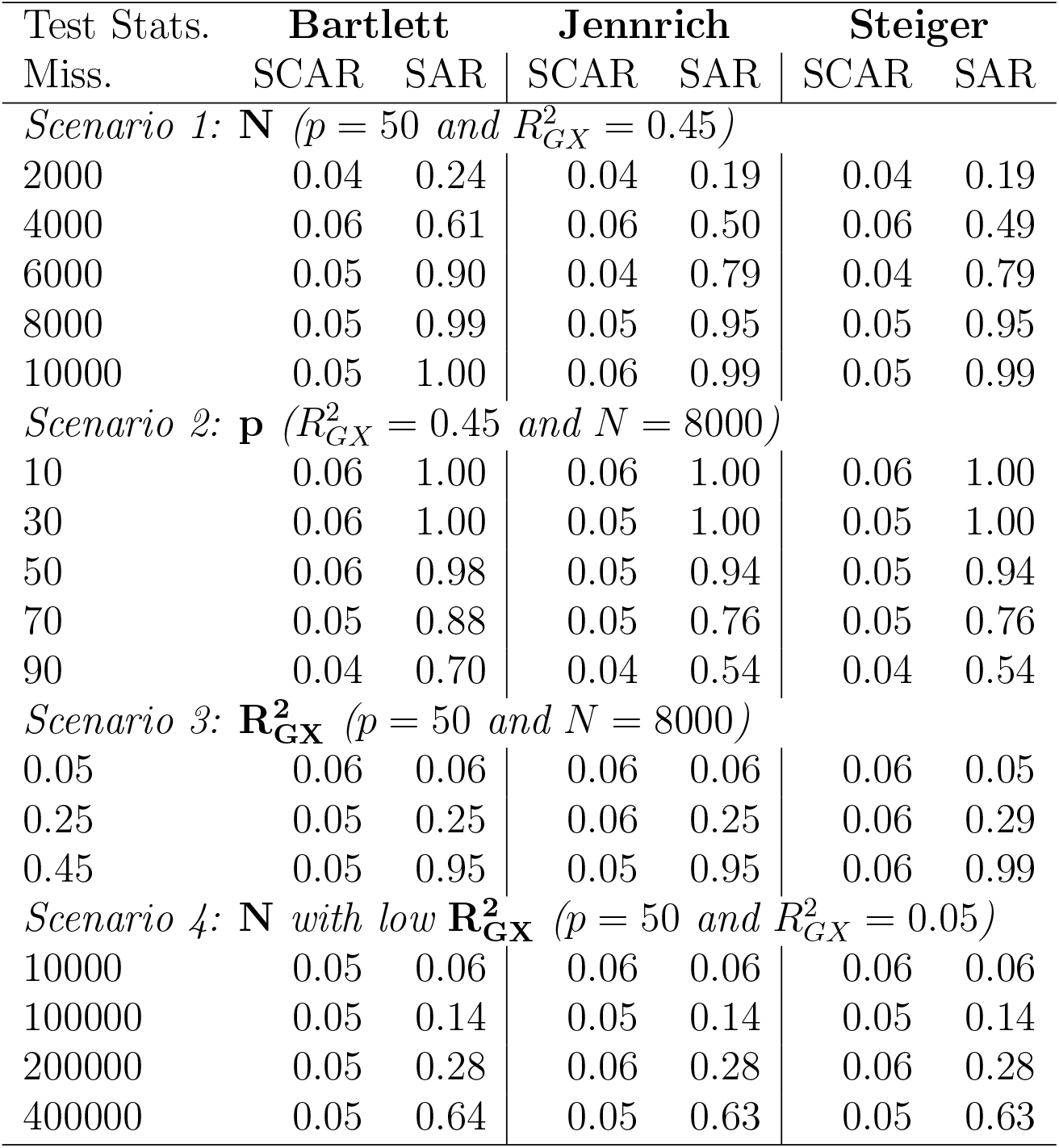
Proportion of tests with p-value *<* 0.05 in 1,000 simulated datasets when testing the identity hypothesis using one sample. Miss., Missingness; SCAR, selection completely at random; SAR, selection at random, conditional on X; *p*, number of *G*s; *N*, sample size; 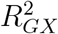, total variance explained by SNPs on X.

We further investigated whether the proportion of tests which correctly reject the null hypothesis when the data are truly SAR improves with greater sample size and fewer *G*s when the variance explained is kept at 5% (Scenario 4). Increasing sample size increases the proportion of tests correctly rejecting the null hypothesis for each of the three methods, however the sample does have to be greater than 400,000 to gain nominal power of 0.8 in our scenario. Reducing the number of *G*s from 90 to 10 only increased the proportion of tests correctly rejecting the null hypothesis in SAR by 0.04 for all three methods (Supplementary Table S1).

### 4.2 Testing the equality of two sample correlation/ co-variance matrices

When testing equality of two sample correlation/covariance matrices under SCAR, all three methods (Steiger, Jennrich and Box’s M) had approximately the nominal 5% T1E (Table 3) in all scenarios. Under SAR, the proportion of the Steiger and Jennrich tests correctly identifying a difference between the two matrices, i.e. with p-value*<* 0.05 was low. Even with a large number of SNPs explaining a high proportion of the variance, the proportion of tests with p-value*<* 0.05 was only slightly above the nominal 0.05 level. Box’s M test correctly rejects the null hypothesis in all 1,000 simulated datasets in all scenarios, except when variance explained by the genetic variants is small.

**Table 3:** Proportion of tests with p-value *<* 0.05 in 1,000 simulated datasets when testing the equality of correlation/covariance matrices from two samples. Miss., Missingness; SCAR, selection completely at random; SAR, selection at random, conditional on X; *p*, number of *G*s; *N*, sample size; 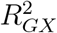, total variance explained by SNPs on X.

| Test Stats. | Box's M |  | Jennrich |  | Steiger |  |
| --- | --- | --- | --- | --- | --- | --- |
| Miss. | SCAR | SAR | SCAR | SAR | SCAR | SAR |
| *Scenario 6: N ($p = 50$ and $R_{GX}^2 = 0.45$)* |  |  |  |  |  |  |
| 5000 | 0.04 | 1.00 | 0.08 | 0.07 | 0.05 | 0.04 |
| 10000 | 0.04 | 1.00 | 0.06 | 0.06 | 0.04 | 0.04 |
| 15000 | 0.04 | 1.00 | 0.06 | 0.05 | 0.05 | 0.04 |
| 20000 | 0.03 | 1.00 | 0.06 | 0.05 | 0.05 | 0.04 |
| *Scenario 7: p ($R_{GX}^2 = 0.45$ and $N = 10000$)* |  |  |  |  |  |  |
| 10 | 0.04 | 1.00 | 0.05 | 0.06 | 0.05 | 0.05 |
| 30 | 0.03 | 1.00 | 0.04 | 0.05 | 0.04 | 0.04 |
| 50 | 0.05 | 1.00 | 0.07 | 0.08 | 0.06 | 0.06 |
| 70 | 0.03 | 1.00 | 0.08 | 0.08 | 0.05 | 0.04 |
| 90 | 0.05 | 1.00 | 0.10 | 0.11 | 0.05 | 0.05 |
| *Scenario 8: $R_{GX}^2$ ($p = 50$ and $N = 10000$)* |  |  |  |  |  |  |
| 0.05 | 0.03 | 0.70 | 0.05 | 0.06 | 0.04 | 0.04 |
| 0.25 | 0.04 | 1.00 | 0.07 | 0.07 | 0.04 | 0.05 |
| 0.45 | 0.04 | 1.00 | 0.07 | 0.05 | 0.05 | 0.04 |
| *Scenario 9: N with low $R_{GX}^2$ ($p = 50$ and $R_{GX}^2 = 0.05$)* |  |  |  |  |  |  |
| 10000 | 0.03 | 0.70 | 0.05 | 0.06 | 0.04 | 0.04 |
| 100000 | 0.04 | 1.00 | 0.06 | 0.06 | 0.06 | 0.06 |
| 200000 | 0.04 | 1.00 | 0.05 | 0.05 | 0.05 | 0.05 |
| 400000 | 0.04 | 1.00 | 0.05 | 0.04 | 0.04 | 0.04 |

The simulated datasets in Scenario 9 have the low variance explained and large sample size that is typically seen in MR study. The number of simulated datasets in which the test correctly rejects the null hypothesis did not increase with sample size for either Steiger or Jennrich’s methods. In all scenarios, Box’s M test has 4% T1E and correctly rejects the null hypothesis in all 1,000 simulated datasets under SAR, except when sample size is small. Fewer *G*s did not improve the number of simulated datasets for which the test correctly rejects the null hypothesis for Steiger or Jennrich’s methods, but did for Box’s M test (Supplementary Table S2).

Both Steiger and Jennrich methods performed poorly for all scenarios when testing equality of the correlation/covariance matrices from two samples. To investigate the reasons for this, we simulated a further three scenarios with larger variance explained (*>*0.45) and lower number of instruments (*<*20), as shown in Supplementary Table S3. Both Jennrich and Steiger test correctly rejected the null hypothesis in 81% and 62% of the simulated datasets once sample size is greater than 10,000, 5 instruments and when variance explained by the genetic variants is 0.9 (Supplementary Table S4).

## 5 Applied data example: alcohol consumption in UK Biobank

Our motivating example of alcohol consumption in UKBB was described in the Introduction. Here we will detail the methods of selection of instruments and quality control excluding SNPs with low MAF, in Hardy-Weinberg disequilibrium and linkage disequilibrium. We then describe how we use the genetic instruments associated with alcohol consumption, BMI, and a random set of genetic variants to explore: 1) selection bias in UK Biobank (by using the one sample methods to compare the correlation matrix between variants within UK Biobank to the identity matrix and 2) selection bias in the analytical sample of those with alcohol consumption measured, by comparing the correlation matrices between those with and without the exposure measured.

### 5.1 Method

A large meta-analysis of GWAS (*N* = 941, 280) identified 99 SNPs associated with weekly alcohol intake (Liu et al., 2019). As negative controls, we randomly selected 99 independent SNPs from across the genome with UKBB (after quality control). As a positive control we selected 82 previously identified body mass index (BMI) SNPs (Locke et al., 2015). The former (random SNPs) are unlikely to be related to selection either into UKBB or by having weekly alcohol intake measured. For the latter (BMI SNPs), there is evidence that obesity levels in UKBB are lower than in the general population (Fry et al., 2017) and thus we expect these SNPs to be correlated in the UKBB sample. We do not expect selection into the sample with alcohol measured to depend on BMI, therefore the correlation matrics between BMI SNPs in those with and without alcohol consumption measured should not differ.

There are in total 391,872 individuals with genotype data in UKBB after restricting the sample to European, unrelated individuals and imputation accuracy greater than 0.8 (Mitchell et al., 2019). 28, 30 and 27 SNPs remained for weekly alcohol intake, random and BMI respectively, when restricted to SNPs that have MAF between 0.1 and 0.5, in Hardy-Weinberg equilibrium and not in linkage disequilibrium (defined as *r*^2^ =0.01, *r*^2^ is estimated from 1000 Genomes European population). In total, the alcohol intake SNPs explain 1% of the variance of alcohol intake, and BMI SNPs explain 2% of the variance of BMI. 45.4% of individuals with genotype data have weekly alcohol intake observed and 99.7% have BMI measured. See Supplementary Table S5, S6 and S7 for phenotypic associations with the three sets of SNPs used. Calculations for linkage disequilibrium structure from 1000 Genomes Project and GWAS associations are from `TwoSampleMR` R package (version 0.5.6).

For testing the identity hypothesis using one sample, the correlation/ covariance from all individuals in UKBB is tested against the identity matrix. Here we are testing for the presence of selection bias in UKBB as a whole. For hypothesis testing for two samples (i.e. equality of the two correlation/ covariance matrices), the matrix derived from the selected sample (those with alcohol intake measured) is tested against the matrix from the non-selected sample (those without alcohol intake measured). Here we are testing for the association of alcohol consumption with having weekly alcohol intake observed. We know from the methodology for UKBB that this selection mechanism is operating, so the aim of this analysis is to see how well the correlation methods detect this known selection mechanism. We have only used Box’s M test for two samples; as demonstrated by the simulations when variance explained is small (Scenario 9) both Jennrich and Steiger test cannot differentiate between missing completely at random and missing at random condition on *X*.

### 5.2 Results

For testing the identity hypothesis in one-sample, shown in Table 4, all the tests demonstrate no correlation between randomly selected SNPs within the full sample. All the tests demonstrate evidence for correlation between alcohol intake and BMI SNPs in the full sample, which suggests that there is selection on alcohol intake and BMI into UKBB related to each of the two sets of SNPs.

**Table 4:** p-value from testing the identity hypothesis in one sample with entire sample of UKBB. Alco. Alcohol intake SNPs and Rand. random SNPs.

| Test/SNPs | Alco. | Rand. | BMI |
| --- | --- | --- | --- |
| Bartlett | <0.001 | 0.1930 | <0.001 |
| Jennrich | <0.001 | 0.1951 | <0.001 |
| Steiger | <0.001 | 0.1951 | <0.001 |

When testing equality of correlation/covariance matrices from two samples, Box’s M test found a difference (p-value *<* 0.001) in covariance matrices between individuals that have weekly alcohol intake measured and unmeasured (i.e. between those with low alcohol frequency and those with higher alcohol frequency). Box’s M test gave no evidence against equality of correlation/covariance matrices between those with and without weekly alcohol data for randomly selected and BMI SNPs (p-value is 0.6487 and 0.1739 respectively).

In conclusion, all the one-sample tests and the two-sample test demonstrated that there is evidence for selection bias due to alcohol intake in UKBB when analysing either the full dataset, or only individuals that had weekly alcohol intake measured.

## 6 Discussion

The Monte Carlo simulation demonstrated that for testing the identity hypothesis using one sample, Steiger, Jennrich and Bartlett tests are able to reject the identity hypothesis in the presence of selection and have nominal type I error rates when selection is completely at random, with large sample size, low number of SNPs and higher proportion of variance explained by the SNPs. When testing equality of correlation/covariance matrices from two samples, Box’s M test is the only approach that is able to detect a difference when selection is conditional on X and has nominal type I error rates when selection is completely at random. We would not recommend Jennrich or Steiger for testing the equality of correlation/covariance matrices from two samples, because as shown by the simulations even with large sample size both test statistics could not distinguish selection completely at random from selection at random. Furthermore, we recommend checking the variance explained by the SNPs, as none of the one-sample tests perform well at detecting selection mechanisms with low variance explained, except with a large sample size.

One previous Monto Carlo simulation study also showed that the proportion of simulations in which the Steiger or Bartlett tests correctly reject the null hypothesis increases with sample size (Brown & Forsythe, 1974). This study also found that Bartlett’s test is sensitive to non-normality (Brown & Forsythe, 1974; Layard, 1973), with an inflated type I error if the variables were from a heavy-tailed error distribution. However, Box’s M test is robust to non-normality (Box, 1953; Layard, 1973). Yang and DeGruttola (2012) have developed a bootstrap version of Bartlett’s test statistic in attempt to reduce inflated type I error and heavy tailed error distribution. Their Monte Carlo simulation used small sample sizes, however, and in GWAS the sample sizes are usually in thousands which will substantially increase computational time to run their approach. Within a GWAS setting, Jennrich’s test statistics have been used for detecting whether shared risk variants of two traits are the result of “subgroup heterogeneity” or “whole-group pleiotropy” (Han et al., 2016). (Han et al., 2016) have also found Jennrich’s test to have low power and modified it by including allele frequency and effect sizes as weights. The test statistics reviewed within our study are designed for low-dimensional setting i.e. the number of variables is less than the sample size. For an extensive review of test statistics for the high-dimensional setting see Zheng et al., 2019.

In our applied example, we identified selection into both UKBB, and the subsample with alcohol intake measured. The design of the questions for UKBB means that only those with higher alcohol consumption had alcohol intake measured. The correlation test used correctly identified that the SNPs for alcohol consumption were correlated differently in the sample with data on alcohol intake than in the sample without. This has implications for any analyses using the alcohol intake measure as either an exposure or an outcome. In this example, the selection mechanism was known (by the design of the survey), and we were using the correlation analysis as a proof of principle. In cases where the selection mechanism is not known, then comparing correlation matrices for genetic variants related to exposure and outcome between those selected and not selected would help to identify whether exposure or outcome cause selection, and therefore the likelihood of selection bias. In the analysis of potential selection into UKBB, the correlation test implied that the SNPs for both alcohol consumption and BMI were correlated within the UKBB sample. Both alcohol consumption and obesity have been shown to be differently distributed in UKBB participants than in the general population (Fry et al., 2017), suggesting that the correlation test is indeed identifying selection mechanisms. Analyses in UKBB with either BMI or alcohol consumption as exposure or outcome may be biased by this selection - for a specific analysis, further examination of the other factors involved in the selection mechanism would be required.

These test statistics require complete-case analysis, as only individuals with an observation for every variable in the correlation matrix will be included in the analysis. Missing SNP data is usually due to poor DNA quality or quantity or technical fault (Pompanon et al., 2005) so the missingness is likely to be completely at random and therefore complete case analysis will not cause bias. However, a complete case analysis would have reduced sample size and thus lower power. Alternatively, if a few SNPs are missing for multiple individuals, then these SNPs could be removed from the analysis. Or, as a third option, the values of the missing SNPs could be imputed, based on the values of SNPs in linkage disequilibrium.

A key advantage of these methods is that they can be used to detect selection (and thus the likelihood for selection bias) even without data on the unselected sample (e.g. by using a correlation matrix from 1000 Genomes Project). In order to identify whether a given analysis is likely to be biased by selection, the selection mechanism must be known (Hughes, Heron, et al., 2019). Usually this is impossible without data on the unselected group. However, if genetic data (or other variables that are known to be independent in the target population) are available, then our proposed use of correlations allows identification of selection using just the observed data.

A researcher with a given analysis question should use knowledge about missing data/selection to identify which selection mechanisms would bias their results. For example, in an MR of alcohol intake on BMI, if selection was related to alcohol intake, this would bias the results Hughes, Davies, et al., 2019. This could be investigated further by taking SNPs identified as related to the exposure and outcome (here, SNPs related to alcohol intake and to BMI) and examining their correlation in the analysis sample. Correlation between these SNPs would indicate that the selection mechanism was caused by those SNPs, and thus by implication caused by the exposure/outcome, and thus that the complete case analysis may be biased.

Another use of these correlation tests is in two-sample MR. The two-sample test for equality of correlation/covariances is one way to examine whether the two samples are from the same underlying population. Even for correlated genetic instruments, two-sample tests can be used to test this assumption.

Selection is not the only possible cause of correlation between genetic variants in a given sample. Assortative mating is another potential explanation. Previous work used genetic correlation to detect assortative mating for genetically predictive traits within the population (Yengo et al., 2018), where under the null hypothesis of random mating, the correlation is zero between alleles on different chromosomes. Detecting assortative mating in MR can be realised through family data, however GWAS family data are sparse and within-family analysis usually lacks power as it requires many mother-father-offspring trios (Howe et al., 2022). For the two-sample test, if assortative mating is the same in two samples being compared (e.g. if they do come from the same population), then this would not cause the correlation matrices from the two samples to differ. In the one-sample test, assortative mating and other forms of selection will not be separable as the one-sample test assumes independence between the SNPs within the underlying population.

In conclusion, we recommend Steiger, Jennrich and Bartlett’s test for one sample and Box’s M test for two sample as sensitivity analyses to identify selection bias in studies where data on genetic variables are available, and to examine the assumption that two samples come from the same underlying population in two-sample MR studies.These hypothesis tests should be examined along side other evidence for selection or assortative mating.These tests will be particularly useful where there are no data on the unselected sample (e.g. for UK Biobank participation).

## Data Availability

The individual-level data that support the findings of this study are available with the permission of the UK Biobank (https://www.ukbiobank.ac.uk). We conducted this study using the UK Biobank resource under an approved data application (ref: 66074).

## Acknowledgments

We thank the reviewers and associate editor for providing comments and suggestions to improve this paper. All authors works in a unit that receives support from the University of Bristol and the UK Medical Research Council (MC UU 00011/1 MC UU 00011/3). This study was conducted under UK Biobank application 66074. *Conflict of Interest*: None declared.

## References

Bartlett, J. W., Harel, O., & Carpenter, J. R. (2015). Asymptotically Unbiased Estimation of Exposure Odds Ratios in Complete Records Logistic Regression. American Journal of Epidemiology, 182 (8), 730–736. https://doi.org/10.1093/aje/kwv114

Bartlett, M. S. (1951). The effect of standardization on a χ 2 approximation in factor analysis. Biometrika, 38 (3/4), 337–344.

Bowden, J., Del Greco M F., Minelli, C., Davey Smith, G., Sheehan, N., & Thompson, J. (2017). A framework for the investigation of pleiotropy in two-sample summary data mendelian randomization. Stat Med, 36 (11), 1783–1802.

Box, G. E. (1949). A general distribution theory for a class of likelihood criteria. Biometrika, 36 (3/4), 317–346.

Box, G. E. (1953). Non-normality and tests on variances. Biometrika, 40 (3/4), 318–335.

Brown, M. B., & Forsythe, A. B. (1974). Robust tests for the equality of variances. Journal of the American Statistical Association, 69 (346), 364–367. Retrieved June 23, 2022, from http://www.jstor.org/stable/2285659

Cai, T. T. (2017). Global testing and large-scale multiple testing for highdimensional covariance structures. Annual Review of Statistics and Its Application, 4, 423–446.

Fry, A., Littlejohns, T. J., Sudlow, C., Doherty, N., Adamska, L., Sprosen, T., Collins, R., & Allen, N. E. (2017). Comparison of sociodemographic and health-related characteristics of UK Biobank participants with those of the general population. American journal of epidemiology, 186 (9), 1026–1034.

Han, B., Pouget, J. G., Slowikowski, K., Stahl, E., Lee, C. H., Diogo, D., Hu, X., Park, Y. R., Kim, E., Gregersen, P. K., et al. (2016). A method to decipher pleiotropy by detecting underlying heterogeneity driven by hidden subgroups applied to autoimmune and neuropsychiatric diseases. Nature genetics, 48 (7), 803–810.

Howe, L. J., Nivard, M. G., Morris, T. T., Hansen, A. F., Rasheed, H., Cho, Y., Chittoor, G., Ahlskog, R., Lind, P. A., Palviainen, T., et al. (2022). Within-sibship genome-wide association analyses decrease bias in estimates of direct genetic effects. Nature genetics, 54 (5), 581– 592.

Hughes, R. A., Davies, N. M., Davey Smith, G., & Tilling, K. (2019). Selection bias when estimating average treatment effects using one-sample instrumental variable analysis. Epidemiology (Cambridge, Mass.), 30 (3), 350.

Hughes, R. A., Heron, J., Sterne, J. A., & Tilling, K. (2019). Accounting for missing data in statistical analyses: Multiple imputation is not always the answer. International journal of epidemiology, 48 (4), 1294–1304.

Jennrich, R. I. (1970). An asymptotic χ2 test for the equality of two correlation matrices. Journal of the American Statistical Association, 65 (330), 904–912.

Larsson, S. C., Burgess, S., Mason, A. M., & Michaelsson, K. (2020). Alcohol consumption and cardiovascular disease: A mendelian randomization study. Circulation: Genomic and Precision Medicine, 13 (3), e002814.

Larzelere, R. E., & Mulaik, S. A. (1977). Single-sample tests for many correlations. Psychological Bulletin, 84 (3), 557.

Layard, M. W. (1973). Robust large-sample tests for homogeneity of variances. Journal of the American Statistical Association, 68 (341), 195– 198.

Liu, M., Jiang, Y., Wedow, R., Li, Y., Brazel, D. M., Chen, F., Datta, G., Davila-Velderrain, J., McGuire, D., Tian, C., et al. (2019). Association studies of up to 1.2 million individuals yield new insights into the genetic etiology of tobacco and alcohol use. Nature genetics, 51 (2), 237–244.

Locke, A. E., Kahali, B., Berndt, S. I., Justice, A. E., Pers, T. H., Day, F. R., Powell, C., Vedantam, S., Buchkovich, M. L., Yang, J., et al. (2015). Genetic studies of body mass index yield new insights for obesity biology. Nature, 518 (7538), 197–206.

Mitchell, R., Hemani, G., Dudding, T., Corbin, L., Harrison, S., & Paternoster, L. (2019). UK Biobank Genetic Data: MRC-IEU Quality Control, version 2. https://doi.org/10.5523/bris.1ovaau5sxunp2cv8rcy88688v

Morris, T. P., White, I. R., & Crowther, M. J. (2019). Using simulation studies to evaluate statistical methods. Statistics in medicine, 38 (11), 2074–2102.

Munafo, M. R., Tilling, K., Taylor, A. E., Evans, D. M., &Davey Smith, G. (2018). Collider scope: When selection bias can substantially influence observed associations. International journal of epidemiology, 47 (1), 226–235.

Neill, J. J., & Dunn, O. J. (1975). Equality of dependent correlation coefficients. Biometrics, 531–543.

Pirastu, N., Cordioli, M., Nandakumar, P., Mignogna, G., Abdellaoui, A., Hollis, B., Kanai, M., Rajagopal, V. M., Parolo, P. D. B., Baya, N., et al. (2021). Genetic analyses identify widespread sex-differential participation bias. Nature Genetics, 53 (5), 663–671.

Pompanon, F., Bonin, A., Bellemain, E., & Taberlet, P. (2005). Genotyping errors: Causes, consequences and solutions. Nature Reviews Genetics, 6 (11), 847–859.

Rothman, K., Greenland, S., & Lash, T. (2008). Modern epidemiology. Wolters Kluwer Health/Lippincott Williams &Wilkins. https://books.google.co.uk/books?id=Z3vjT9ALxHUC

Stamatakis, E., Owen, K. B., Shepherd, L., Drayton, B., Hamer, M., & Bauman, A. E. (2021). Is cohort representativeness passe? poststratified associations of lifestyle risk factors with mortality in the UK Biobank. Epidemiology (Cambridge, Mass.), 32 (2), 179.

Steiger, J. H. (1980). Testing pattern hypotheses on correlation matrices: Alternative statistics and some empirical results. Multivariate Behavioral Research, 15 (3), 335–352.

Yang, Y., & DeGruttola, V. (2012). Resampling-based methods in single and multiple testing for equality of covariance/correlation matrices. The international journal of biostatistics, 8 (1).

Yengo, L., Robinson, M. R., Keller, M. C., Kemper, K. E., Yang, Y., Trzaskowski, M., Gratten, J., Turley, P., Cesarini, D., Benjamin, D. J., et al. (2018). Imprint of assortative mating on the human genome. Nature human behaviour, 2 (12), 948–954.

Zheng, S., Cheng, G., Guo, J., & Zhu, H. (2019). Test for high dimensional correlation matrices. Annals of statistics, 47 (5), 2887.

