## Supplementary Materials for "Use of genetic correlations to examine selection bias"

Table S1: Proportion of tests with p-value $< 0.05$ in 1,000 simulated datasets with testing the identity hypothesis using one-sample. $R^2_{GX}$ is 0.05 and sample size of 8,000. Miss., Missingness; $p$, number of $G$s; $N$, sample size; $R^2_{GX}$, total variance explained by SNPs on X.

| Test Stats. | **Steiger** | | **Jennrich** | | **Bartlett** | |
|---|---|---|---|---|---|---|
| Miss. | SCAR | SAR | SCAR | SAR | SCAR | SAR |
| *Scenario 5: number of $G$s with low $R^2_{GX}$* | | | | | | |
| 10 | 0.06 | 0.08 | 0.06 | 0.08 | 0.06 | 0.08 |
| 30 | 0.06 | 0.05 | 0.06 | 0.05 | 0.06 | 0.06 |
| 70 | 0.04 | 0.07 | 0.05 | 0.07 | 0.04 | 0.07 |
| 90 | 0.06 | 0.05 | 0.06 | 0.05 | 0.06 | 0.04 |

Table S2: Proportion of tests with p-value $< 0.05$ in 1,000 simulated datasets when testing the equality of correlation/covariance matrices from two-samples. $R^2_{GX}$ is 0.05 and sample size of 10,000. Miss., Missingness; $p$, number of $G$s; $N$, sample size; $R^2_{GX}$, total variance explained by SNPs on X.

| Test Stats. | **Steiger** | | **Jennrich** | | **Box's M** | |
|---|---|---|---|---|---|---|
| Miss. | SCAR | SAR | SCAR | SAR | SCAR | SAR |
| *Scenario 10: number of $G$s with low $R^2_{GX}$* | | | | | | |
| 10 | 0.05 | 0.04 | 0.05 | 0.04 | 0.04 | 0.99 |
| 30 | 0.04 | 0.05 | 0.04 | 0.06 | 0.03 | 0.92 |
| 50 | 0.06 | 0.06 | 0.07 | 0.08 | 0.05 | 0.68 |
| 70 | 0.05 | 0.04 | 0.08 | 0.07 | 0.03 | 0.44 |
| 90 | 0.05 | 0.05 | 0.10 | 0.12 | 0.05 | 0.32 |

Table S3: Summary of simulation scenarios. $p$, number of $G$s; $N$, sample size; $R^2_{GX}$, total variance explained by $G$s on X.

| **Scenario** | $p$ | $R^2_{GX}$ | $N$ |
|---|---|---|---|
| 11 | 5 | 0.9 | 2000-10000 |
| 12 | 5-20 | 0.9 | 10000 |
| 13 | 5 | 0.45-0.9 | 10000 |

Table S4: Proportion of tests with p-value $< 0.05$ in 1,000 simulated datasets when testing the equality of correlation/covariance matrices from two-samples. Miss., Missingness; $p$, number of $G$s; $N$, sample size; $R^2_{GX}$, total variance explained by SNPs on X.

| Test Stats. | **Steiger** | | **Jennrich** | | **Box's M** | |
|---|---|---|---|---|---|---|
| Miss. | SCAR | SAR | SCAR | SAR | SCAR | SAR |
| *Scenario 11: sample size ($p = 5$ and $R^2_{GX} = 0.9$)* | | | | | | |
| 2000 | 0.06 | 0.12 | 0.06 | 0.33 | 0.05 | 1.00 |
| 4000 | 0.05 | 0.26 | 0.06 | 0.55 | 0.05 | 1.00 |
| 6000 | 0.06 | 0.43 | 0.06 | 0.67 | 0.04 | 1.00 |
| 8000 | 0.03 | 0.52 | 0.03 | 0.78 | 0.03 | 1.00 |
| 10000 | 0.04 | 0.59 | 0.05 | 0.80 | 0.05 | 1.00 |
| *Scenario 12: number of Gs ($R^2_{GX} = 0.9$ and $N = 10000$)* | | | | | | |
| 5 | 0.06 | 0.63 | 0.06 | 0.82 | 0.05 | 1.00 |
| 10 | 0.04 | 0.08 | 0.04 | 0.35 | 0.04 | 1.00 |
| 15 | 0.05 | 0.06 | 0.06 | 0.20 | 0.05 | 1.00 |
| 20 | 0.06 | 0.04 | 0.06 | 0.16 | 0.04 | 1.00 |
| *Scenario 13: variance explained ($p = 5$ and $N = 10000$)* | | | | | | |
| 0.45 | 0.06 | 0.09 | 0.06 | 0.14 | 0.05 | 1.00 |
| 0.6 | 0.05 | 0.22 | 0.05 | 0.34 | 0.04 | 1.00 |
| 0.75 | 0.04 | 0.40 | 0.04 | 0.61 | 0.03 | 1.00 |
| 0.9 | 0.05 | 0.62 | 0.05 | 0.81 | 0.04 | 1.00 |

Table S5: Associations between weekly alcohol intake SNPs and weekly alcohol intake in UK Biobank

| SNP | Chr | Pos | EA | NEA | Beta | SE | p-val |
|---|---|---|---|---|---|---|---|
| rs10506274 | 12 | 81601464 | T | G | 0.011 | 0.003 | 5.00E-04 |
| rs1104608 | 16 | 73912588 | C | G | 0.017 | 0.003 | 1.70E-08 |
| rs1123285 | 14 | 57274519 | G | C | 0.016 | 0.003 | 1.40E-06 |
| rs113443718 | 16 | 29892184 | A | G | 0.027 | 0.003 | 6.80E-17 |
| rs11739827 | 5 | 166803321 | T | G | 0.015 | 0.003 | 4.50E-07 |
| rs11940694 | 4 | 39414993 | G | A | -0.044 | 0.003 | 1.00E-44 |
| rs12088813 | 1 | 66407700 | C | A | 0.009 | 0.003 | 1.20E-02 |
| rs1260326 | 2 | 27730940 | C | T | -0.051 | 0.003 | 3.30E-60 |
| rs12795042 | 11 | 133658168 | C | A | 0.012 | 0.003 | 1.40E-04 |
| rs12907323 | 15 | 86796012 | G | A | -0.008 | 0.003 | 9.50E-03 |
| rs13250583 | 8 | 20949917 | T | C | 0.012 | 0.004 | 8.80E-04 |
| rs13383034 | 2 | 45155276 | T | C | -0.020 | 0.003 | 5.80E-10 |
| rs2165670 | 4 | 100286085 | A | G | -0.047 | 0.005 | 3.30E-21 |
| rs2764771 | 16 | 20013793 | A | G | -0.026 | 0.003 | 9.80E-15 |
| rs281379 | 19 | 49214274 | A | G | -0.019 | 0.003 | 4.10E-10 |
| rs2854334 | 17 | 29715500 | G | A | -0.016 | 0.003 | 5.30E-07 |
| rs28601761 | 8 | 126500031 | G | C | -0.013 | 0.003 | 3.60E-05 |
| rs3748034 | 4 | 3446091 | T | G | 0.025 | 0.004 | 1.40E-08 |
| rs378421 | 16 | 28754684 | A | G | 0.034 | 0.003 | 1.80E-27 |
| rs3809162 | 12 | 54674235 | G | A | -0.027 | 0.003 | 6.30E-18 |
| rs4842786 | 12 | 92170791 | A | G | 0.012 | 0.003 | 1.20E-04 |
| rs500321 | 13 | 27124360 | T | A | 0.008 | 0.003 | 2.80E-02 |
| rs55932213 | 9 | 108755622 | G | A | -0.012 | 0.004 | 6.20E-04 |
| rs56030824 | 11 | 47397353 | A | G | 0.022 | 0.003 | 9.90E-12 |
| rs56337305 | 2 | 225475560 | C | T | 0.007 | 0.003 | 1.90E-02 |
| rs62250685 | 3 | 85457240 | G | A | 0.021 | 0.003 | 3.20E-11 |
| rs6460047 | 7 | 73042443 | C | T | -0.020 | 0.004 | 1.30E-07 |
| rs6951574 | 7 | 153489744 | C | T | -0.017 | 0.003 | 2.30E-08 |
| rs7185555 | 16 | 69131281 | C | G | 0.017 | 0.004 | 4.10E-05 |
| rs9838144 | 3 | 131576287 | C | G | 0.021 | 0.004 | 4.10E-08 |

Table S6: Associations between randomly selected SNPs and weekly alcohol intake in UK Biobank

| SNP | Chr | Pos | EA | NEA | Beta | SE | p-val |
|---|---|---|---|---|---|---|---|
| rs11773926 | 8 | 24433531 | G | A | 0.009 | 0.003 | 0.0087 |
| rs72771839 | 5 | 87763957 | A | G | 0.015 | 0.004 | 0.0002 |
| rs9991896 | 4 | 6444182 | T | G | 0.007 | 0.004 | 0.0570 |
| rs2144880 | 20 | 59285178 | T | C | 0.003 | 0.003 | 0.3500 |
| rs808233 | 14 | 58390589 | T | C | -0.001 | 0.003 | 0.8000 |
| rs10138601 | 14 | 34296324 | T | C | -0.005 | 0.004 | 0.2500 |
| rs12483418 | 21 | 15442979 | A | G | -0.003 | 0.004 | 0.4500 |
| rs378363 | 9 | 9020223 | C | T | 0.000 | 0.004 | 0.9700 |
| rs17335397 | 8 | 77230117 | C | T | 0.003 | 0.003 | 0.3200 |
| rs2447862 | 5 | 52178971 | C | T | -0.000 | 0.004 | 0.9900 |
| rs2827550 | 21 | 23885063 | G | A | 0.002 | 0.004 | 0.5800 |
| rs12983479 | 19 | 36291664 | A | G | -0.004 | 0.005 | 0.3800 |
| rs61969470 | 13 | 86443326 | G | A | -0.001 | 0.004 | 0.9000 |
| rs10905706 | 10 | 10313455 | G | A | 0.005 | 0.003 | 0.1100 |
| rs2223859 | 20 | 2393386 | A | G | -0.004 | 0.003 | 0.1700 |
| rs6070194 | 20 | 56211410 | C | T | 0.001 | 0.004 | 0.8400 |
| rs56870642 | 18 | 1666022 | A | G | 0.003 | 0.003 | 0.2800 |
| rs34471402 | 9 | 103262534 | C | T | 0.001 | 0.004 | 0.8600 |
| rs10060818 | 5 | 67859256 | G | A | 0.019 | 0.005 | 0.0001 |
| rs1610395 | 10 | 61240403 | C | T | -0.002 | 0.003 | 0.5500 |
| rs10878056 | 12 | 63975707 | A | G | -0.002 | 0.003 | 0.5100 |
| rs45617131 | 2 | 223140055 | G | A | 0.005 | 0.004 | 0.2400 |
| rs759183 | 7 | 13295262 | A | G | 0.003 | 0.004 | 0.4700 |
| rs820390 | 17 | 73747796 | T | A | -0.001 | 0.004 | 0.7400 |
| rs2834710 | 21 | 36350221 | T | C | -0.001 | 0.003 | 0.8300 |
| rs7559399 | 2 | 208911363 | T | C | -0.005 | 0.003 | 0.1300 |
| rs464047 | 5 | 66161048 | G | A | 0.007 | 0.003 | 0.0260 |
| rs11651622 | 17 | 29137155 | G | A | -0.011 | 0.005 | 0.0150 |

Table S7: Associations between BMI SNPs and BMI in UK Biobank

| SNP | Chr | Pos | EA | NEA | Beta | SE | p-val |
|---|---|---|---|---|---|---|---|
| rs10182181 | 2 | 25150296 | G | A | 0.033 | 0.002 | 3.60E-64 |
| rs10938397 | 4 | 45182527 | G | A | 0.029 | 0.002 | 6.90E-48 |
| rs10968576 | 9 | 28414339 | G | A | 0.024 | 0.002 | 2.50E-30 |
| rs11030104 | 11 | 27684517 | G | A | -0.038 | 0.002 | 1.30E-53 |
| rs1167827 | 7 | 75163169 | G | A | 0.021 | 0.002 | 1.80E-25 |
| rs12446632 | 16 | 19935389 | A | G | -0.031 | 0.003 | 3.90E-27 |
| rs12885454 | 14 | 29736838 | A | C | -0.017 | 0.002 | 7.50E-16 |
| rs12940622 | 17 | 78615571 | A | G | -0.018 | 0.002 | 9.60E-19 |
| rs13021737 | 2 | 632348 | G | A | 0.055 | 0.003 | 8.30E-99 |
| rs1516725 | 3 | 185824004 | C | T | 0.032 | 0.003 | 1.80E-29 |
| rs1558902 | 16 | 53803574 | A | T | 0.073 | 0.002 | 4.90E-291 |
| rs16951275 | 15 | 68077168 | C | T | -0.029 | 0.002 | 1.50E-35 |
| rs17405819 | 8 | 76806584 | C | T | -0.020 | 0.002 | 3.70E-21 |
| rs2033529 | 6 | 40348653 | G | A | 0.022 | 0.002 | 1.20E-23 |
| rs2033732 | 8 | 85079709 | C | T | 0.011 | 0.002 | 2.40E-06 |
| rs2112347 | 5 | 75015242 | G | T | -0.027 | 0.002 | 2.70E-40 |
| rs2121279 | 2 | 143043285 | T | C | 0.011 | 0.003 | 2.60E-04 |
| rs2207139 | 6 | 50845490 | G | A | 0.038 | 0.003 | 1.00E-47 |
| rs2245368 | 7 | 76608143 | T | C | -0.022 | 0.003 | 9.80E-17 |
| rs2820292 | 1 | 201784287 | C | A | 0.017 | 0.002 | 7.80E-18 |
| rs3810291 | 19 | 47569003 | A | G | 0.028 | 0.002 | 8.90E-39 |
| rs3888190 | 16 | 28889486 | A | C | 0.027 | 0.002 | 1.20E-39 |
| rs543874 | 1 | 177889480 | G | A | 0.049 | 0.002 | 2.70E-91 |
| rs6567160 | 18 | 57829135 | C | T | 0.054 | 0.002 | 2.30E-118 |
| rs7138803 | 12 | 50247468 | A | G | 0.029 | 0.002 | 9.30E-46 |
| rs9400239 | 6 | 108977663 | C | T | 0.016 | 0.002 | 6.00E-14 |
| rs9925964 | 16 | 31129895 | G | A | -0.022 | 0.002 | 9.60E-27 |
